# Characterizing social contact patterns in rural and urban Mozambique: the GlobalMix study, 2021-2022

**DOI:** 10.1101/2024.06.04.24308064

**Authors:** Moses C. Kiti, Charfudin Sacoor, Obianuju G. Aguolu, Alana Zelaya, Holin Chen, Sara S. Kim, Nilzio Cavele, Edgar Jamisse, Corssino Tchavana, Americo Jose, Ivalda Macicame, Orvalho Joaquim, Noureen Ahmed, Carol Y. Liu, Inci Yildirim, Kristin Nelson, Samuel M. Jenness, Herberth Maldonado, Momin Kazi, Rajan Srinivasan, Venkata R. Mohan, Alessia Melegaro, Fauzia Malik, Azucena Bardaji, Saad B. Omer, Ben Lopman

## Abstract

1.

**Introduction:** There are few sources of empirical social contact data from resource-poor settings thus limiting the development of contextual mathematical models of disease transmission and control.

**Methods:** We collected and analyzed cross-sectional survey data from rural and urban sites in Mozambique. Participants, including infants, were recruited. They reported retrospectively, in a paper diary, individuals with whom they had a co-located physical or conversation contact, as well as their age, sex, relationship, frequency, and duration of the contact. We compared vaccine effects (VE) by parameterizing transmission models using empirical and synthetic contact rates.

**Results:** 1363 participants recruited between April 2021 and April 2022 reported a mean of 8.3 (95% CI 8.0–8.6) contacts per person on day 1. Mean contact rates were higher in the rural compared to urban site (9.8 [9.4–10.2 vs 6.8 [6.5–7.1], p<0.01), respectively. Participants aged ≤18 years were the main drivers of higher physical contacts. In the model, we report higher VE in the rural site when comparing empirical to synthetic contact matrices (32% vs 29%, respectively), and lower corresponding VE in urban site (32% vs 35%). These effects were prominent in the younger (0-9 years old) and older (60+ years) individuals.

**Conclusion:** Our work suggests differences in contact rates and patterns between rural and urban sites in Mozambique, with corresponding differences in vaccine effects on an infectious pathogen. We also demonstrate the utility of empirical data in infectious disease modelling for high-burden, low-income settings.

## 2. Background

Human social contact patterns drive the transmission of pathogens that spread through proximity to infectious individuals. Data on social contact patterns are critical to understand “who-contacts-whom” and infer who-acquires-infection-from-whom, providing insight on potential control measures such as physical distancing and vaccination. Underlying the patterns of contact are demographic, sociocultural, and economic determinants which vary within and across regions resulting to corresponding variation in contact patterns. Unfortunately, such critical data are not as widely available in low-and-middle-income countries (LMICs), including Mozambique [1], as they are in high-income countries (HICs) [2]. Where data are available, they were collected across e.g., rural-urban divide [3–6] or informal settlement [7,8] limiting the representativeness of the data. More recent studies in LMICs have incorporated innovative methods to obtain data from hard-to-reach populations especially children and illiterate adults by either using shadows [3,9] interviewer-led questionnaires [4,5], or wireless proximity sensors [10,11]. In the absence of locally collected data, simulated contact rates for LMIC populations may also be derived by projecting empirical data collected from HICs (e.g., POLYMOD data) and scaled using local demographic patterns [12]. However, these extrapolations likely mischaracterize contact patterns in important ways when they differ for reasons aside from demographics.

During the early phase of the COVID-19 pandemic, it became clear the extent to which human contact patterns could be modified, and at essentially a global scale [13–17] In the absence of vaccines or pharmaceutical interventions, populations responded by “physical distancing”, i.e., drastically reducing the number and riskiness of contacts. Work contacts and those out of the home decreased and masking increased. Again, however, relatively little data were collected from LMICs, limiting our ability to quantify these changes and use such data to develop models of interventions.

Starting 2021, during the COVID-19 pandemic, we launched the “GlobalMix Study” to collect social contact data from four LMICs: Mozambique, Guatemala, India, and Pakistan. Across all four countries, we are collecting data from selected rural and urban areas using standardized questionnaires and methods that were customized for each context considering social and cultural norms [18]. In this paper, we present the methods and results from Mozambique for which we have complete datasets.

## 3. Methods

### 3.1 Study objectives

The main aim of this study was to characterize the age and location patterns of social contact and mixing with respect to directly transmitted infections in rural and urban sites of Mozambique. Then, to examine the impact of using these contextual data, we simulated the transmission of a hypothetical respiratory virus and effects of vaccination in a model using contact data generated from this study (henceforth called empirical data) and compared to empirically constructed contact data (henceforth called synthetic data).

### 3.2 Study design

This was a cross-sectional study conducted from March 2021 through to April 2022. Our data collection period coincided with active SARS-CoV-2 transmission in Mozambique [19]. The rural site was in Manhiça District where the Manhiça Health and Research Centre (CISM) runs the Manhiça Health and Demographic Surveillance System (Manhiça HDSS) [20]. The urban site was in Maputo City where the Polana-Caniço Health and Research Centre (CISPOC) of the National Institute of Health of Mozambique runs the Polana-Caniço HDSS [21]. Prior to collecting the social contact data, we held 25 focus group discussions and 40 cognitive interviews with community members drawn from the two sites. We aimed to understand the determinants of human interaction at the study sites, and explore the perceptions, acceptability, and utility of paper diaries for data collection. We also wanted to get community buy-in and get good practice recommendations on our research implementation process.

Complete details of the sample size, data collection tools and procedures, and data analysis methods have been described in our protocol [18]. Briefly, in Mozambique, we aimed to collect data from 630 individuals per site equally stratified into 10 age groups: 0-5 months, 6-11 months, 1-4, 5-9, 10-14, 15-19, 20-20, 30-39, 40-59 and 60+ years, resulting in 63 participants per age group. The data collection was divided into three phases: enrollment, diary-keeping, and exit interview. Individuals were randomly selected by age and sex from the HDSS registers. Each participant filled in their social contacts over two days. We defined a social contact (henceforth called contact) as a two-way face-to-face encounter that involved either i) physical touch involving a skin-to-skin touch or over clothes, or 2) non-physical, whereby two or more individuals had a conversation interaction only while standing within arm’s length of each other and with no physical barrier between them. Additional qualitative questions included difficulty encountered during filling the diary and perceived social behavior and mobility change during the study period compared to periods before the COVID-19 outbreak. Contact data, the enrollment and exit interview were captured electronically by fieldworkers via REDCap [22] forms coded in portable electronic tablets. The diary codebook in English is available in our GitHub repository (See **10. Availability of data and** materials section). All children <10 years old and illiterate individuals ≥10 years old selected or were assigned a shadow to record contacts on behalf of the participant. In addition, the shadows were instructed to be discrete and did not need to follow the participant all day.

### 3.3 Data analysis

#### 3.3.1 Characteristics of contact patterns

We estimated the mean contact rates per person with bootstrapped 95% confidence intervals (CI) and median contact rates per person with inter-quartile range (IQR) over two days and for day 1 only. Assuming *x_ij_* represents the total number of contacts between participants in age group *i* and contacts in age group 1 *j*, then the mean number of reported contacts (*m_ij_*) was calculated as *x_ij_/n_i_*, where *n_i_* is the study population in group *i*.

The mean contact rates were stratified by site, then further by age, sex, day of the week (weekday vs weekend), type of contact (physical or conversation only), household membership (household member vs non-household member), occupation/ daily activity, and whether the participant reported symptoms of acute respiratory infections (ARI) or acute gastroenteritis (AGE) within the 14 days prior to survey. We used the Wilcoxon rank sum test to assess the difference between median contact rates within the sites for each covariate, and between the rural and urban sites.

Lastly, we computed age-stratified contact matrices to quantify the interactions between age groups. We adjusted the contact matrices to account for reciprocity, assuming that the total to, i.e., *m_i,j_* = *m_j,i_* [6]. We present the age-specific contact matrix using data from day 1 only using the revised formula ((*m_i,j_* + *m_j,i_*)/(*n_i_* + *n_j_*)) which does not adjust for Mozambique age-specific population size.

#### 3.3.2 Characterizing location-specific proximity contact exposures

We compared close, individually recorded diary contacts (close contacts) to co-location with others but without direct interaction (proximity contacts) collected using the place-use surveys. We describe the number of unique visits locations including other person’s home, street, market/shop, transport/hub, agricultural field, school, work, place of worship, well, playground or other and the distribution of time spent at each place. We then compared the number of proximity contacts per participant at each location type to the total close contacts per participant at the same location type we examined for differences in patterns between the urban and rural site.

#### 3.3.3 Transmission and vaccine effects modeling

To explore the utility of the empirical contact patterns, we compared rural and urban respiratory virus transmission models parameterized with empirical data to those parameterized using synthetic contact data modelled using Mozambique demographic data and POLYMOD data [12]. We built a deterministic susceptible-infectious-recovered (SIR) model with a vaccine conferring protection against infection. We computed the mean number of contacts by re-classifying the participants into six 10-year age classes from 0–59 years and one age group for individuals aged 60 years or older for compatibility with the synthetic data. We weighted the empirical contact rates using 2021 rural and urban population distribution data and adjusted for reciprocity using the *socialmixr* package. We modeled vaccination as ‘leaky,’ providing partial protection for those vaccinated (50% vaccine coverage, 50% effectiveness), duration of illness as 7 days and fixed the basic reproduction number at 2.5. We calculated the attack rate for no vaccine (AR_0_) and vaccine (AR_v_) scenarios separately for rural and urban site and present the overall vaccine effect (VE) calculated as the percent reduction of cases in the presence versus absence of vaccine, i.e.,

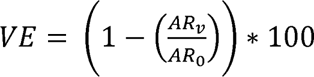. We used the *EpiModel* package to run all transmission models. All analysis was done in R v4.3.2.

### 3.6 Ethical considerations

This study was approved by CISM Internal Scientific Committee (CCI/03/2020), the Internal Ethical Review Board (initial approval CIBS-CISM/011/2020), the Emory University Institutional Review Board (approval number 00105630), and Yale University (reliance agreement approval number 2000026911). Field staff were responsible for obtaining written informed consent from each participant prior to any data collection. Adults (≥18 years) provided written consent, children (13–17 years) required personal written assent and parental consent, while minors (<13 years) only required parental consent.

## 4. Results

### 4.1 Baseline characteristics of participants

We approached 1693 residents and retained a total of 1363 (81% overall participation rate) participants across both sites. We report equal participation rate in the rural (676/800, 85%) and urban (687/893, 86%) site. We exceeded our target sample size by 103 participants particularly in those aged 40–59 years. Of the 1363 participants, 666 (49%) were female equally distributed by site. By site, there was no major difference in number of participants recruited by age, sex, and school enrollment status (**Table 1**).

**Table 1.** Characteristics of study participants in rural and urban sites in Mozambique.

|  |  | **Overall**<br>**N = 1363 (%)** | **Rural**<br>**N = 676 (%)** | **Urban**<br>**N = 687 (%)** |
| --- | --- | --- | --- | --- |
| **Sex<sup>1</sup>** |  |  |  |  |
|  | Female | 666 (49) | 332 (49) | 334 (49) |
|  | Male | 696 (51) | 343 (51) | 353 (51) |
| **Participant age** |  |  |  |  |
|  | <6mo | 128 (9) | 62 (9) | 66 (10) |
|  | 6-11mo | 146 (11) | 82 (12) | 64 (9) |
|  | 1-4y | 135 (10) | 63 (9) | 72 (10) |
|  | 5-9y | 122 (9) | 64 (9) | 58 (8) |
|  | 10-14y | 125 (9) | 61 (8) | 64 (9) |
|  | 15-19y | 124 (9) | 64 (9) | 60 (9) |
|  | 20-29y | 125 (9) | 64 (9) | 61 (9) |
|  | 30-39y | 125 (9) | 64 (9) | 61 (9) |
|  | 40-59y | 209 (15) | 89 (13) | 120 (17) |
|  | 60+y | 124 (9) | 63 (9) | 61 (9) |
| **Able to read and write** |  |  |  |  |
|  | Yes | 701 (51) | 293 (43) | 408 (59) |
| **Currently enrolled in school** |  |  |  |  |
|  | Yes | 368 (28) | 173 (26) | 195 (29) |
| **Occupation/ daily activity<sup>2</sup>** |  |  |  |  |
|  | Child | 274 (23) | 144 (24) | 130 (22) |
|  | Unemployed | 162 (14) | 97 (16) | 65 (11) |
|  | Student | 324 (27) | 153 (26) | 171 (29) |
|  | Homemaker | 33 (3) | 9 (2) | 24 (4) |
|  | Casual laborer | 78 (7) | 22 (4) | 56 (9) |
|  | Farmer | 70 (6) | 64 (11) | 6 (1) |
|  | Businessperson | 66 (6) | 7 (1) | 59 (10) |
|  | Office worker | 83 (7) | 27 (5) | 56 (9) |
|  | Retired | 20 (2) | 5 (1) | 15 (3) |
|  | Other | 74 (6) | 58 (10) | 14 (2) |
| **Regular mask use** |  |  |  |  |
|  | Yes | 942 (69) | 435 (64) | 507 (74) |
| **Acute gastroenteritis** |  |  |  |  |
|  | Diarrhea/ vomiting | 27 (2) | 15 (2) | 12 (2) |
| **Acute respiratory infection** |  |  |  |  |
|  | >1 symptom | 233 (17) | 122 (18) | 111 (16) |
| **Who filled the diary?** |  |  |  |  |
|  | Self | 665 (49) | 361 (54) | 304 (44) |
|  | By fieldworker | 698 (51) | 315 (46) | 383 (56) |

<sup>1</sup> 2 participants did not report their sex.
<sup>2</sup> 179 participants did not report their Occupation/ daily activity

The mean household size was 5.5 (range 1–18) and 5.7 (range 1–20) in the rural and urban sites, respectively. Overall, 379 (45%) households had 4-6 members and 4 (5%) had one resident. When we omit children, students and the unemployed, the most common reported occupations in the urban site were small business owners and vendors classified as businesspeople (16%, 59/366), office workers (15%, 56/366), and casual laborers (15%, n=56/366) while farmers comprised 16% (64/394) in the rural site. 942 (69%) of participants reported wearing a mask inside or outside the house, with no difference by site. About one-fifth (233) of participants reported having >1 ARI symptom and 26 (2%) reported at least one AGE symptom.

About half (51%, n=701) of the participants were able to read and write. The majority (88%, n=1200) of the participants said that they reported all contacts. However, 51% (698/1363) required assistance from a fieldworker to fill in the diary at the end of the two days (rural 43% vs urban 56%). Generally, all children <5 years (n=409) had a family member as a shadow, and out of these 243 (50%) required additional assistance from the fieldworker. Among other ages, there was no difference in proportion of those requiring a shadow/ help from fieldworker compared to no help, apart from 15–19 (33% [41/124]) and 60+ year-olds (60%, [75/124]). Eight participants reported testing positive for SARS-CoV-2, all of whom reported either going to quarantine (government facility, n=5) or self-isolation at home (n=3).

### 4.2 Contact patterns

Participants reported a total of 17674 contacts over two days, with 41% unique contacts (n=3904 day 1 only and n=3250 day 2 only) and 59% (n=10304) reported on both days (repeat contacts). Participants reported an overall mean of 13.1 (95% CI 12.6–13.5) contacts over two days (**Table 2**). We observe a significant difference in the mean number of contacts reported on day 1 compared to day 2 (8.3 [8.0–8.6] vs 5.5 [5.3–5.7], respectively, paired t-test p<0.01). Since diary completion dates were randomly assigned, the actual mean contacts should not vary between the first and second date of diary completion. Therefore, we believe that the observed difference is a result of reporting bias resulting from participant fatigue; from here henceforth we report the mean and median number of contacts on day 1 only. The rural mean contact rate was significantly higher than the urban rate (9.8 [9.4–10.2] vs 6.8 [6.5–7.1], p<0.01) (**Figure 1** A & B). Contact rates were higher in rural areas for each age group (Figure 1 C). While rural mean number of contacts with non-household members was significantly higher that contacts with household members (6.6 vs 3.9, p<0.01), there was marginal difference in the urban site (4.2 vs 3.6, p=0.46). Corresponding median values for day 2 by site are available in **Supplementary table 1**.

**Figure 1.**
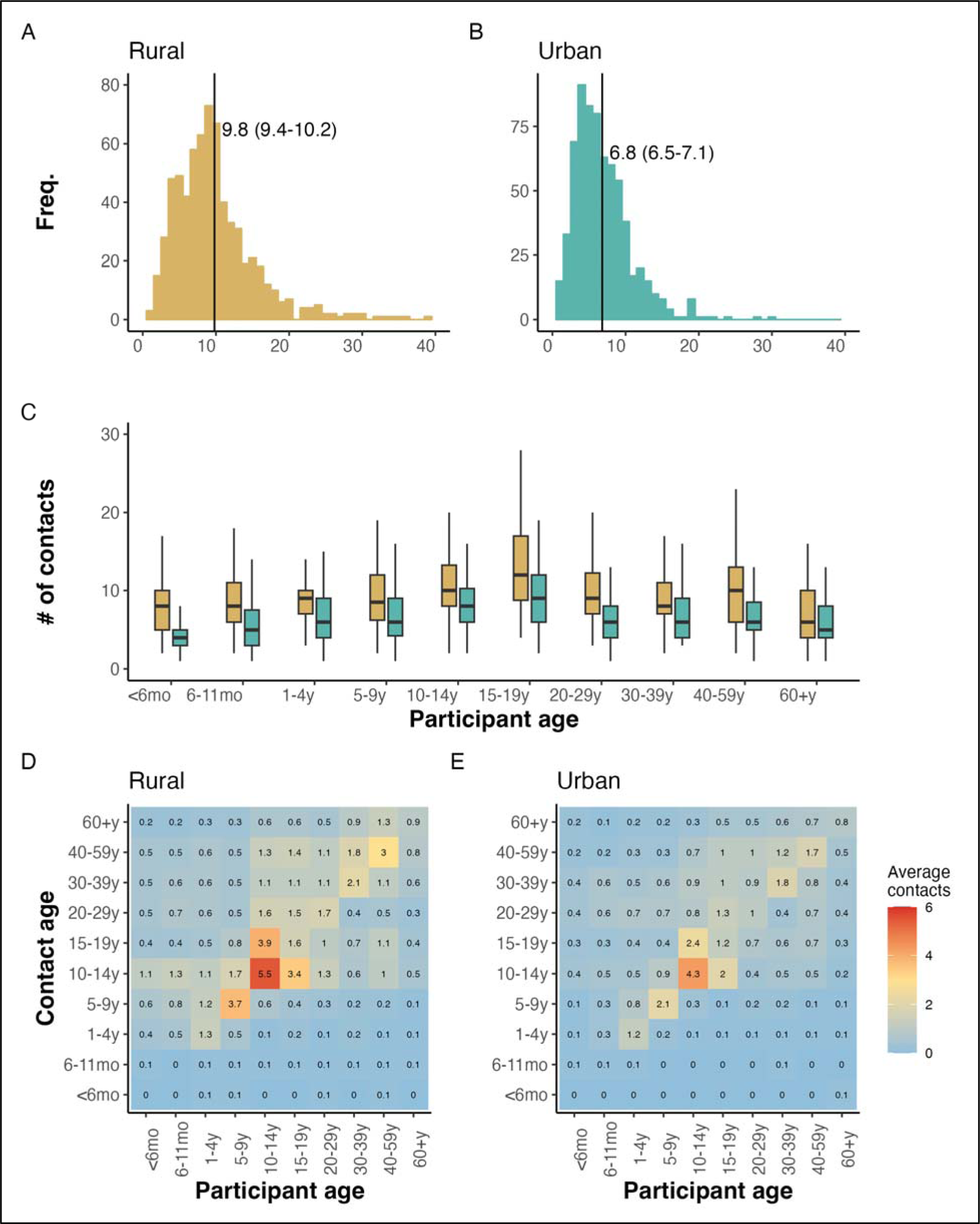
Distribution patterns of number of contacts in rural and urban Mozambique. Panels A and B show the density distribution of the number of contacts per person in the rural and urban site, respectively, with the mean and 95% CI shown by the black vertical line. Panel C shows boxplots of the distribution of number of contacts by site. The middle line shows the median number of contacts and the lower and upper line in the boxplot shows the lower and upper quartile of the distribution, respectively. Panels D and E show the age specific contact matrices depicting the average mean number of individuals in age-group (y-axis) with whom a

**Table 2.**
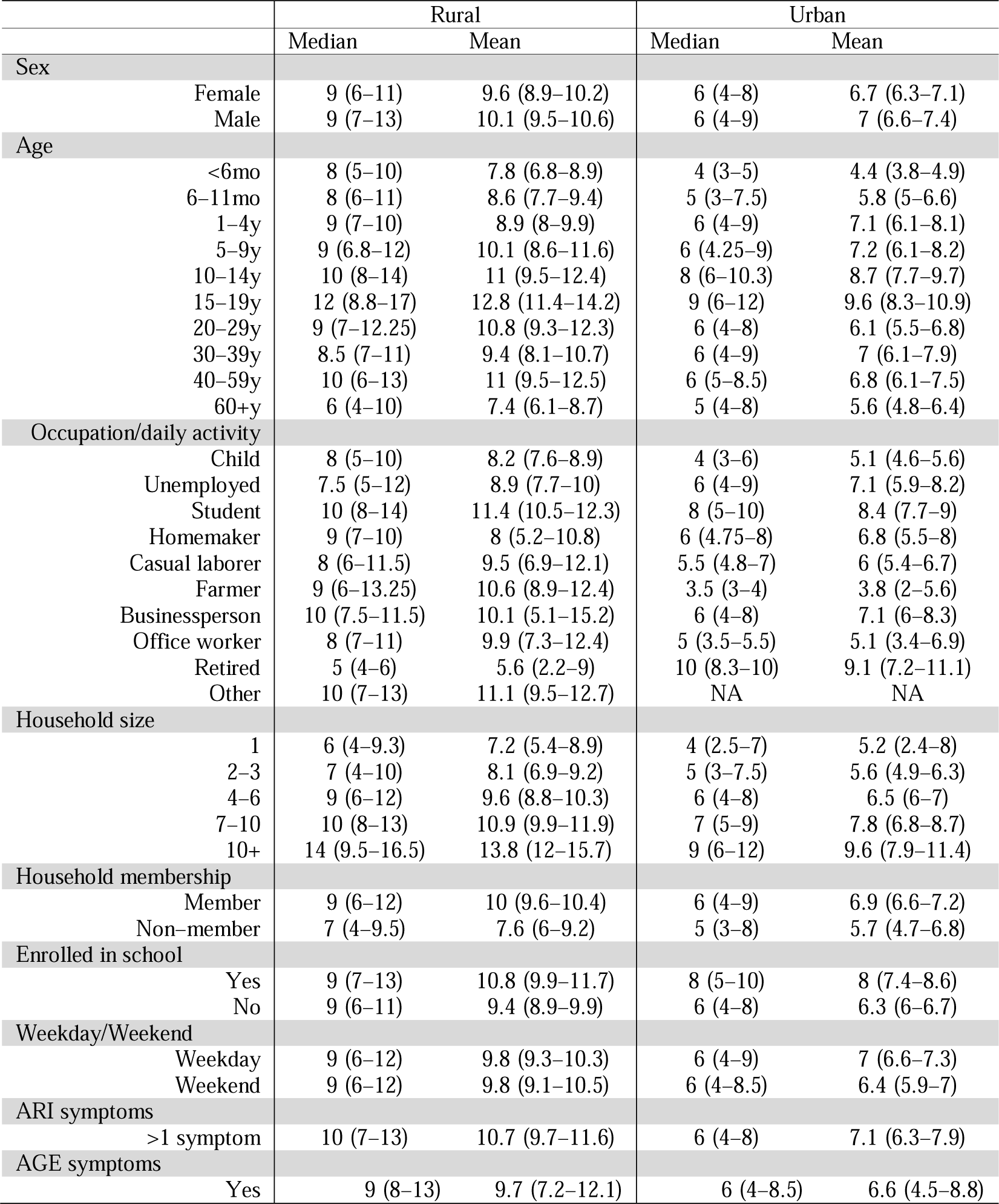
Median (IQR) and mean (bootstrapped 95% CI) number of contacts on day 1 only.

Physical contacts were, on average, more numerous than conversation only contacts in both rural (6.7 vs 4.9, p<0.01) and urban areas (5.3 vs 3.3, p<0.01). Participants aged ≤18 years were the main drivers of the higher physical contacts. Out of all participants, 803 (59%) reported having the same number of social contacts compared to periods before the COVID-19 pandemic. From these, urban participants reported either significantly fewer (n=238, mean = 5.9 [95% CI 5.5– 6.3]) or more (n=28, 11.7 [9.3–14.1]) mean number of contacts compared to those who reported no change (n=410, mean = 7.0 (6.7–7.4)). In the rural site, 74 (11%) of the participants reported more mean contacts than usual (12.6, 95% CI 10.9–14.3) and this was significantly different from those who reported either no change (n=384, mean = 9.6 [9.1–10.1]) or fewer contacts (n=215, mean = 9.2 [8.5–9.9]).

#### 4.2.1 Contact matrices

The patterns of age-specific social contacts are generally similar when comparing the urban and rural sites (**Figure 1** panel D and E). The urban matrix, however, suggests lower mean number of contacts across all ages compared to the rural site. Participants aged 5-14 years (school-going children) and older working adults aged 30–59 years in both rural and urban areas reported higher assortative (with same age) mean contacts. Older school goers aged 15–19 years also reported on average, high number of contacts with 10–14-year-olds in both sites. Another peak in mean number of contacts is observed between 30–39- and 40–59-year-olds, driven mostly by conversation only contacts (**Supplementary figure 2** panels (C) and (D)). In both sites, few interactions are reported between the other ages and little to no contacts are reported with infants. Despite low mean contacts reported by infants, the mean number of contacts between them and other ages generally increased with age to peak at 10–14 years in rural and 30–39 years among urban dwellers.

#### 4.2.2 Patterns of contact by location

In both urban and rural areas, the estimated number of co-located individuals by far exceeded the number of contacts reported by participants. Overall, participants reported a mean of 26.9 (95% CI 23.5–30.3) proximity contacts compared to 6.8 (6.5–7.1) close contacts in the urban site, and 23.1 (20.3–25.9) proximity contacts and 9.8 (9.4–10.2) close contacts in the rural site. The three locations with highest mean number of contacts were places of worship, schools, and transport hubs (**Table 3**). In addition, rural participants were more likely (n=752 visits, [48%]) to visit other homes compared to urban participants (n=288 visits [29%]). Despite overall numbers being similar, the locations where contact occurred was meaningfully different between urban and rural sites.

**Table 3.** Number of times that participants reported visiting a location (N) and the median (IQR) number of people reported per location on day 1.

| Place visited | Rural |  | Urban |  |
| --- | --- | --- | --- | --- |
|  | N (%) | Median (IQR) | N (%) | Median (IQR) |
| Other home | 752 (48) | 4 (3–6) | 261 (29) | 4 (3–7) |
| Street | 369 (23) | 2 (1–5) | 342 (37) | 4 (3–9) |
| Market/Shop | 92 (6) | 6 (3–12) | 77 (8) | 10 (3–30) |
| Transport/Hub | 90 (5) | 16 (11–18) | 60 (6) | 18 (12–20) |
| Agricultural field | 104 (7) | 3 (1.8–6.3) | 8 (1) | 5.5 (1–6.8) |
| School | 62 (3) | 25 (15–25.8) | 63 (6) | 30 (24–33.5) |
| Work | 45 (2) | 6 (4–20) | 54 (6) | 6 (3–10) |
| Place of worship | 26 (2) | 20 (10.3–30) | 23 (2) | 30 (17.5–45) |
| Well | 17 (1) | 4 (2–6) | 1 | 7 |
| Playground | 1 | 15 | 1 | 13 |
| Other | 45 (3) | 10 (5–23) | 47 | 7 (3.5–18) |

#### 4.2.3 Sensitivity of transmission model disease dynamics to our empirical contact matrices

To recap, we restructured the age-specific matrices presented in **Figure 1** D and E into 7 age groups (see Methods) depicted in **Figure 2** panels A and B. Assortative contacts in empirical data were highest among 10–19-year-olds (higher in rural compared to urban) compared to synthetic values for 0–9-year-olds (panel C, data available from [12]).

**Figure 2.**
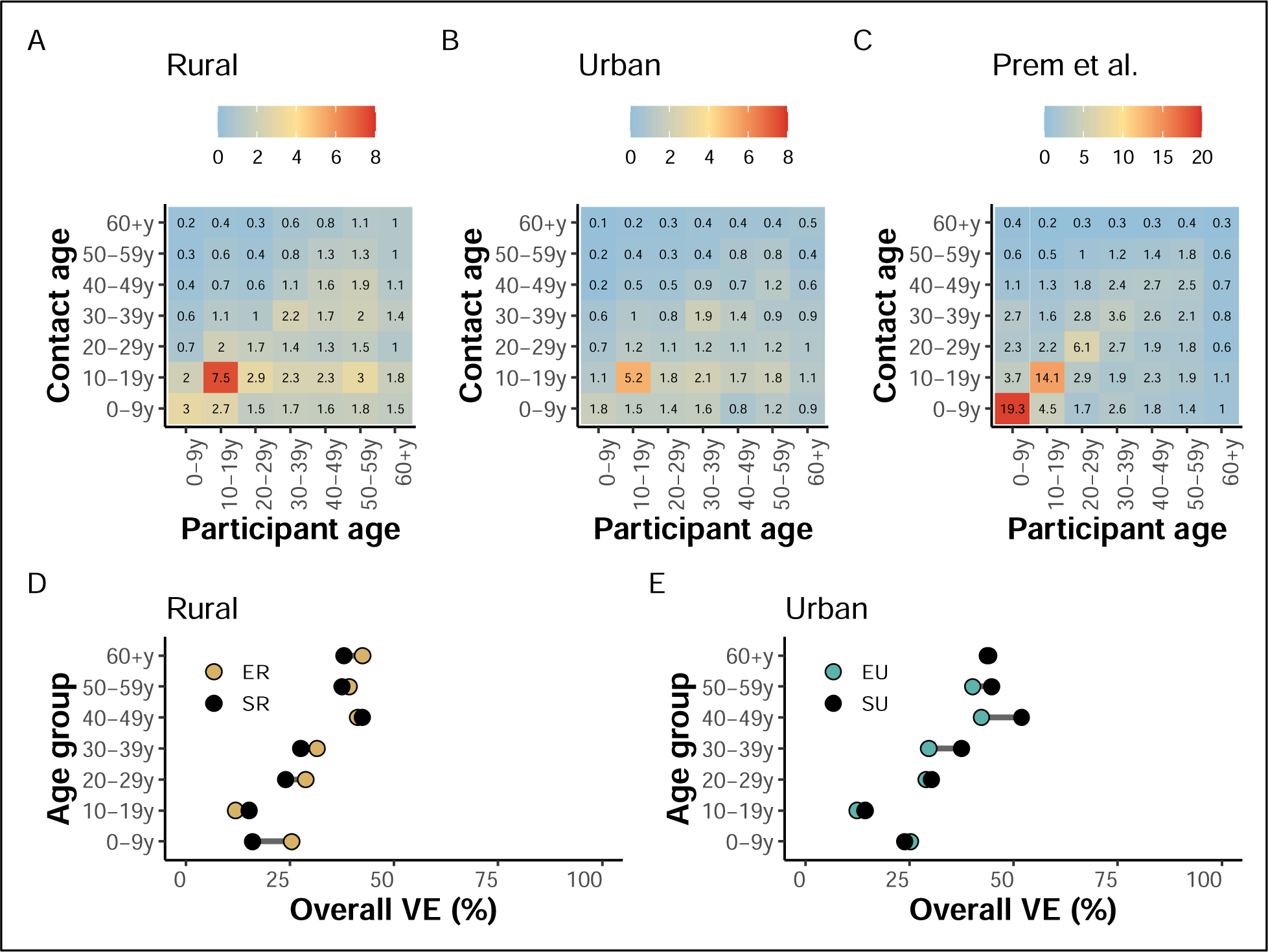
Contact matrices and modeled age-specific vaccine effects (VE) Panels A and B show rural and urban contact matrices from our empirical data, respectively, while panel C shows the synthetic contact matrix derived from Mozambique-specific demographic data by Prem et al [12]. Panels D and E shows the overall vaccine effects (VE) of a respiratory infection comparing synthetic and empirical contact rates in the rural and urban site, respectively. Legend key: ER=Empirical Rural, SR=Synthetic Rural, EU=Empirical Urban, SU=Synthetic Urban.

In the rural and urban site, the values of the empirical overall vaccine effects (VE) overlapped in most ages. However, synthetic VE were marginally higher in 10–19 and 40–49-year-olds in the rural site (**Figure 2** D), and higher in all ages (particularly adults aged 30–59-year-olds) apart from children aged 0–9-year old (**Figure 2** E) in the urban site. Specifically, we report higher attack rates in unvaccinated (AR) compared to vaccinated (ARv) individuals (AR: 94%, ARv: 77%) and lower overall vaccine effects (18%) for 0–9-year-olds compared to attack rates using empirical data (rural AR: 84%, rural ARv, 62%, rural vaccine effects: 26%; urban AR: 84%, urban ARv: 62%, urban vaccine effects: 26%). Additionally, synthetically contacts among 60+ years old were underestimated compared to empirical values, producing notably lower attack rates among this age group (Synthetic AR: 49%, Synthetic ARv: 31%; Rural AR: 40%, Rural ARv: 24%; Urban AR: 36%, Urban ARv: 21%).

## 5. Discussion

We present results from a two-day cross-sectional study aiming to quantify social contact rates among residents of a rural and urban site in Mozambique during the COVID-19 pandemic. We first engaged with the local community to get their views on the suitability and acceptability of our tools and study procedures. We made several key observations. First, we used the qualitative outcomes to modify the format and content of the paper diaries to make them more user friendly (see additional details in **Supplementary text 1**). Second, in our sample population, participants from the rural site had significantly higher average number of contacts compared to the urban site. Third, in each site, reported mean contacts increased with age to peak at school-going children/ teenagers aged 15-19 years, and mean contacts were higher among adults (>18 years) compared to children <5 years. Fourth, mixing was assortative (increased frequency of contacts within the same ages) among school-going children, and less pronounced inter-generational mixing particularly in the urban site. Finally, we quantify the importance of using empirical contact data to model disease dynamics and interventions. In model simulations of an emerging respiratory pathogen, we find meaningfully different attack rates and vaccine impact among both child and elderly groups when using our local data compared with widely used contact matrices modelled from other settings.

In the earlier phases of the COVID-19 pandemic, Mozambique adopted physical distancing policies including school closures and restrictions on social gatherings, but with no express requirement to stay at home [23]. This was similar to measures implemented globally to reduce transmission of SARS-CoV-2. Few countries in sub-Saharan Africa e.g., South Africa [24], Kenya [3,5], Zimbabwe [4], Uganda [9], and Somaliland [8] had existing empirical social contact data collected in various settings prior to 2020. There are sparse data on contact patterns during the pandemic from LMICs (examples are from Kenya [7] and Malawi [6]) compared to high income countries such as the United Kingdom and Europe [17] and USA [16]. Longitudinal data during the pandemic would have been critical for a better understanding of the transmission pathways of the novel virus in LMICs taking account of demographic, social and economic contexts, and provide insights on how to enhance NPI use in place and identify priority groups for immunization once vaccines became available albeit in limited supply and use in LMICs.

Compared to pandemic periods in Kenya [7] and Malawi [6], we observed lower mean number of contacts in Mozambique but a higher number of contacts reported by participants in the rural compared to the urban site. However, we interpret this with care since data from Kenya and Malawi were collected from high-density settlements where individuals may have been unable to fully adhere to physical distancing mainly due to economic reasons. The Government of Mozambique periodically revised physical distancing policies to curb the spread of SARS-CoV-2 [25], and we also think that the measures were not strictly adhered to particularly among school-going children and working adults.

We implemented several innovations in the GlobalMix study. First, we collected contact data from participants over two consecutive days. The average number of contacts stratified across different covariates of interest remained constant over two days, suggesting the stability of participant’s recall as well as the number and nature of contacts made over multiple days. The stability of contact networks across days has been previously demonstrated in rural households in Kenya and Malawi using autonomous methods that minimize recall bias [10,11]. A second innovation of our study was an estimation of group proximity contacts at locations frequently visited by participants. It is important to note that participants report almost four times the number of proximity contacts compared to detailed individually reported contacts. This suggests the potential to substantially underestimate the number of interactions that could lead to transmission events particularly in highly mobile age groups congregating in social venues.

Finally, our transmission model simulation demonstrates the importance of contextual empirical social contact data. Although advanced methods for projecting social contact patterns onto regions without data exist (e.g., [12]), we found that age-specific infection attack rates from a model using empirical contact data differed meaningfully compared to a model parameterized with synthetic contact rates. Importantly, we found the largest differences in attack rates (comparing vaccinated versus unvaccinated individuals) resulted in increased vaccine impact in the youngest (0-9 years) age group, who often represent the most vulnerable group. These findings were consistent with a Uganda model where use of local contact pattern data resulted in larger epidemics in young children and smaller epidemics in adults aged >35 years compared to using UK-based contact data [9]. It is also notable that we observed distinct contact patterns resulting in divergent model results for the Mozambique rural and urban sites. This highlights the impact of sub-national differences in contact patterns resulting in variation in disease dynamics. Such insights are not possible with widely used contact data which are generally available at the national level.

## 6. Conclusion

We present empirical results of a cross-sectional study quantifying rates and patterns of human social contacts relevant for the spread of directly transmitted infections in rural and urban areas of Mozambique. We demonstrate the possibility of collecting high quality social contact data from resource-poor settings thus reducing reliance on synthetic data modelled from high-income countries. Finally, we demonstrate the potential advantages of empirical compared to synthetic data in a transmission and vaccine control model and advocate for the use of contextual data in similar studies. Nevertheless, some questions remain including if social contact patterns changed in this setting as non-pharmaceutical interventions were relaxed. Finally, we are completing data collection from three other LMICs and we will make all our data collection tools, data, and analysis scripts findable, accessible, interoperable, and reusable (FAIR).

## Data Availability

ar

## 8. Acknowledgements

We acknowledge all the community members and local authorities who have supported and/or participated in data collection. We also thank all the researchers and field staff involved in this work, both from Manhiça and Polana Caniço HDSS. Our special thanks are extended to the National Institute of Health of Mozambique (INS) and the CISPOC Directorate, who willingly participated in the research by including the urban site of the Polana-Caniço HDSS.

## 10. Availability of data and materials

All data produced in the present study are available on our GitHub repository: https://github.com/lopmanlab/globalmix-mozambique-aim1. We also include the survey codebooks in English and Portuguese, as well as the Portuguese version of the diary to portray the age group images used in the study.

## 11. Authors contribution

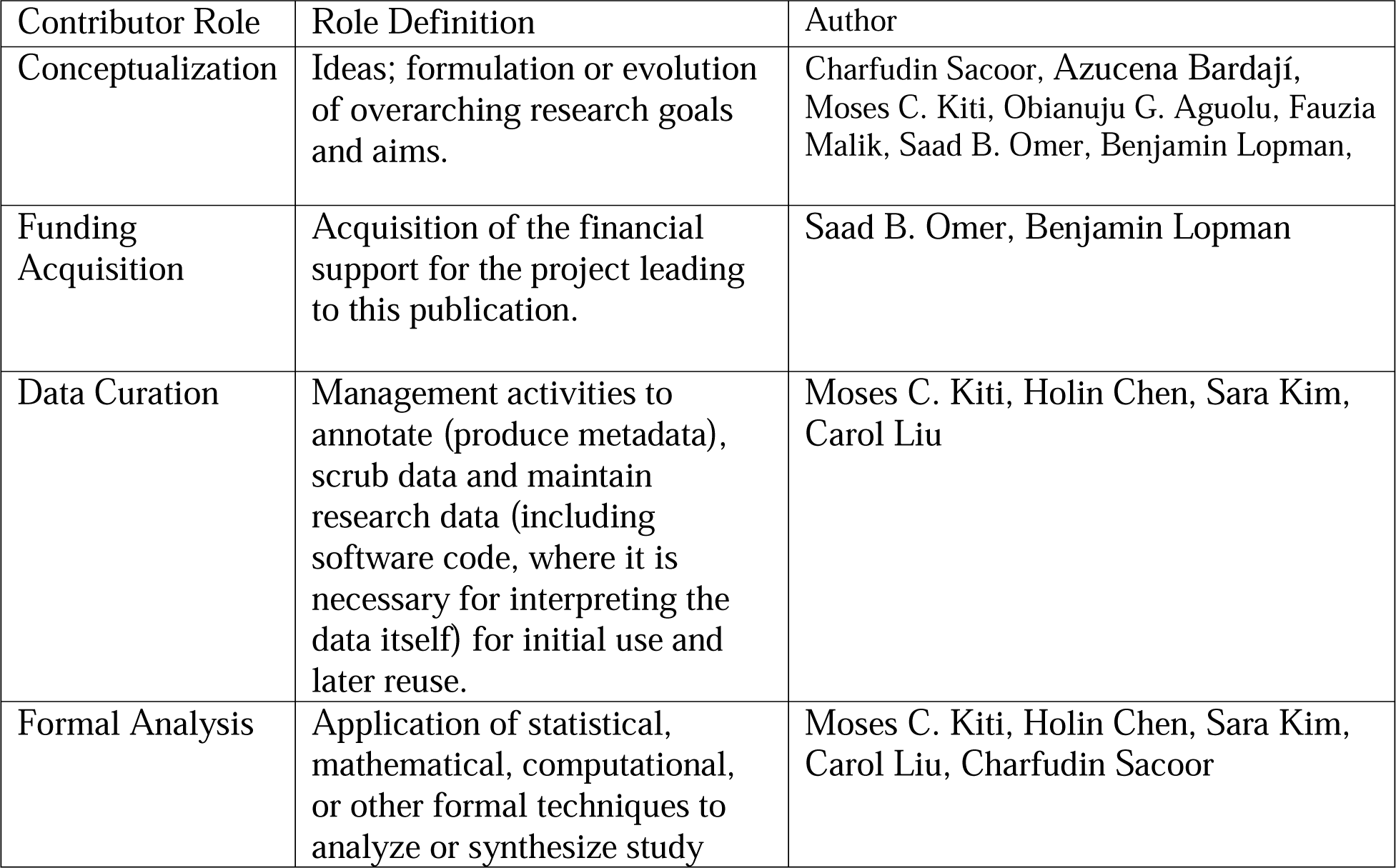
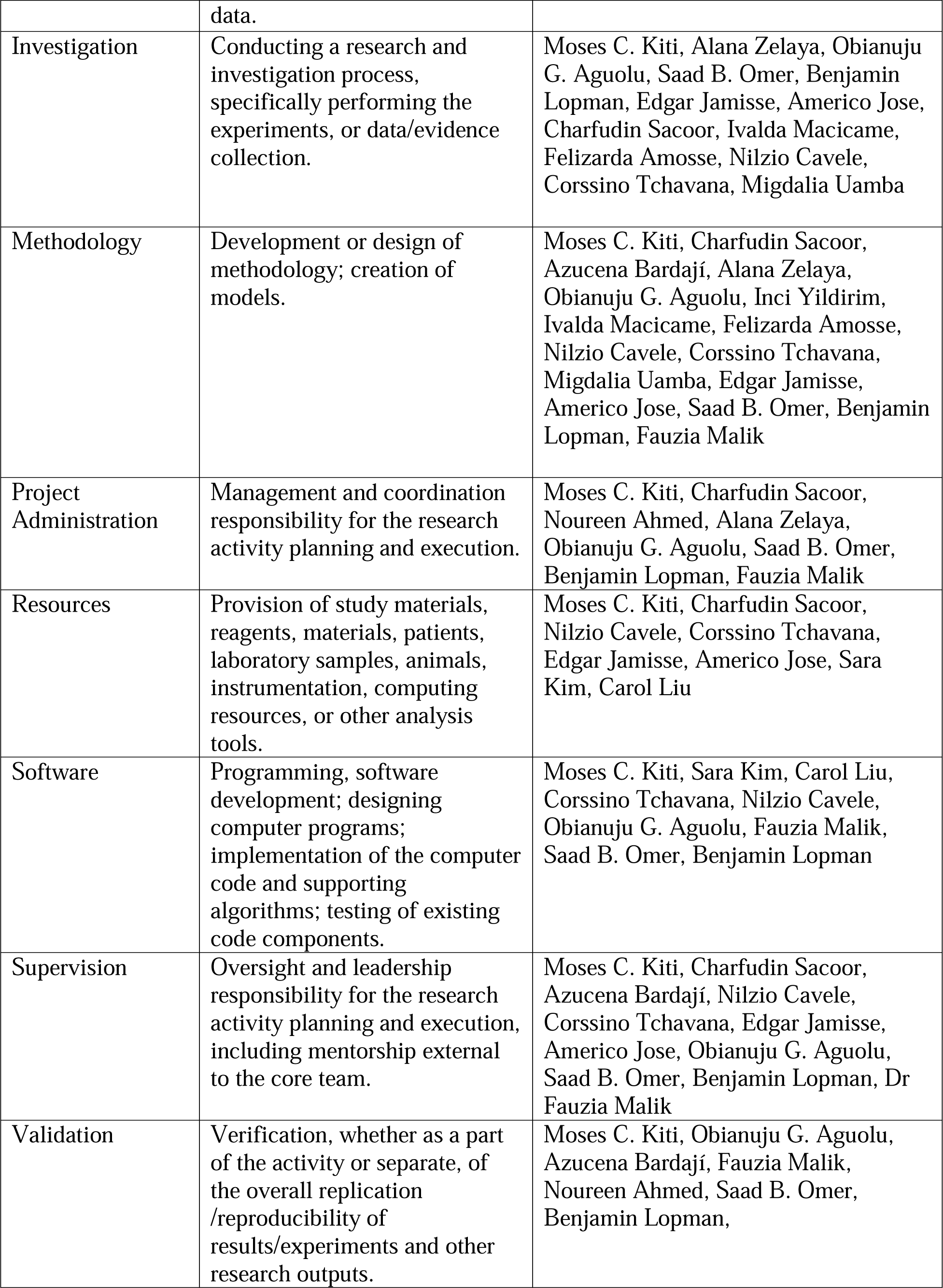
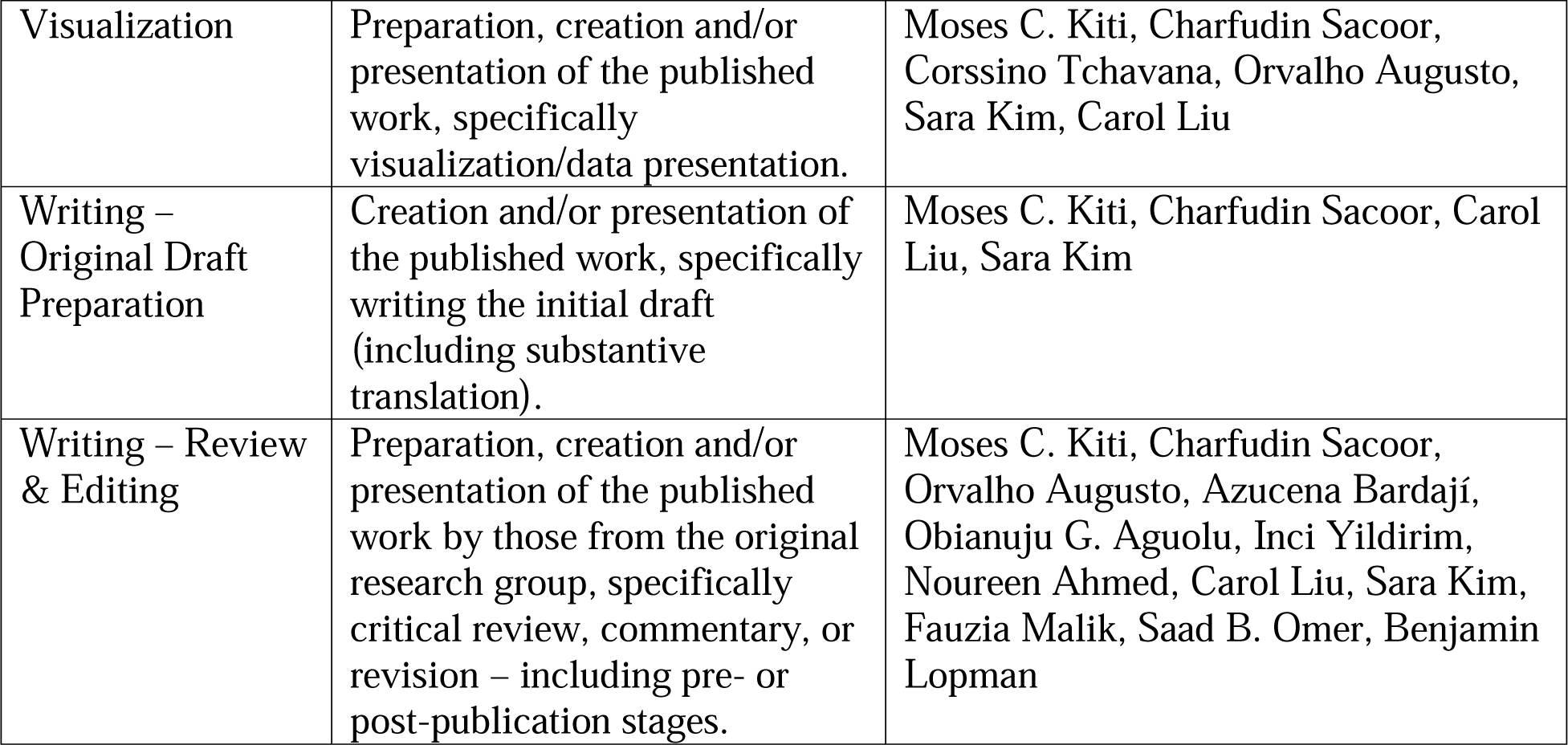

## 13. Supplementary Information

**Supplementary text 1**: Details of fieldwork, challenges experienced, and practical solutions applied.

This was the first time that self-report paper diaries were being used in the Manhiça and Polana Canhiço HDSS. It is important to note the logistic and technical challenges we experienced during the conduct of our study, and our mitigation measures, to better inform future studies in similar settings. Following extensive consultations with community members, we used paper diaries with contextualized questions and customized visual aids to guide age selection of contacts, including those met for the first time. During implementation, adult males were hard to find since most left their houses early for work, market sales and to tend to their farms. Absenteeism was also reported in a similar study in Kenya [3] and the authors replaced the loss-to-follow-up individuals with others of similar demographic characteristics. Due to the pandemic, both HDSS databases had not been updated and we resorted to door-to-door recruitment for under-5-year-olds and later expanding the study catchment area to additional HDSS blocks in the urban site. During the rainy season, remote areas in the rural site were inaccessible thus hampering recruitment and data collection exercises. Once recruited, some participants were unavailable to present their diary data during the exit visits, and almost half of diaries in both sites were partly filled in, thus requiring assistance from our field staff to complete the diary. Eventually, we resorted to recruiting children from mothers attending health clinics, visits to workplaces for adult males, and following up participants through phone calls. The lapse in time between diary data collection period and exit survey could have contributed to recall bias and it was not possible to ascertain the magnitude and direction of the recall bias. Despite this, we had a good buy in from the community due to our extensive community engagement conducted before and during the study, which also resulted in higher participation rates in general than previously observed in other sub-Saharan African settings. Furthermore, the data was collected during periods of active transmission of SARS-CoV-2 in the community, including certain times when certain restrictive measures were in place, such as the closure of schools and parks, limitations on the number of employees at the workplace and passengers on public transportation, social distancing, and other measures [23]. This context likely caused some disruption, given the contingency scenario at the time, and may have influenced the characteristics of social contact patterns due to the ongoing pandemic.

**Supplementary table 1:** Median (IQR) rural contacts on day 2.

|  |  | Rural | Urban |
| --- | --- | --- | --- |
| Sex |  |  |  |
|  | Female | 5 (3–9) | 5 (3–8) |
|  | Male | 8 (5–11) | 6 (4–8) |
| Age |  |  |  |
|  | <6mo | 5 (3–7) | 2 (1–4) |
|  | 6–11mo | 5 (3–9) | 3.5 (2–5.8) |
|  | 1–4y | 5 (3–8) | 5 (3–7) |
|  | 5–9y | 6 (4–9) | 4 (2–5) |
|  | 10–14y | 6 (3–9) | 4 (3–7) |
|  | 15–19y | 7 (4–12) | 6 (3–7) |
|  | 20–29y | 6 (4–9) | 4 (3–5) |
|  | 30–39y | 5 (3–10) | 4 (2.8–6) |
|  | 40–59y | 5 (3–9) | 3 (2–4) |
|  | 60+y | 4 (2–7) | 3 (1.3–4.8) |
| Occupation/ daily activity |  |  |  |
|  | Child | 5 (3–8) | 3 (2–5) |
|  | Unemployed | 4.5 (2–8) | 3.5 (2–5.3) |
|  | Student | 6 (4–10) | 4 (3–7) |
|  | Homemaker | 3 (2–6.5) | 2 (2–3) |
|  | Casual laborer | 6.5 (2.8–10.3) | 3.5 (2–4.8) |
|  | Farmer | 5 (2–7) | 9 (5–9) |
|  | Businessperson | 9 (4–9.5) | 3 (2–4) |
|  | Office worker | 5 (3.5–11) | 4 (2–5) |
|  | Retired | 7 (5.8–7) | 1.5 (1–2.8) |
|  | Other | 5.5 (4–10) | 5 (4–6) |
| Household size |  |  |  |
|  | 1 | 6 (2–10) | 5.5 (4–8) |
|  | 2–3 | 5.5 (3–9) | 4 (2–6) |
|  | 4–6 | 5 (3–8.8) | 4 (2–5) |
|  | 7–10 | 5 (3–7.8) | 3 (2–5) |
|  | 10+ | 5 (3–6.8) | 2 (2–5) |
| Enrolled in school |  |  |  |
|  | Yes | 6 (3–10) | 4 (2–7) |
|  | No | 5 (3–9) | 3 (2–5) |
| Weekday/Weekend |  |  |  |
|  | Weekday | 5 (3–9) | 4 (2–6) |
|  | Weekend | 5 (3–9) | 4 (2–5) |
| ARI symptoms |  |  |  |
|  | No symptom | 5 (3–9) | 4 (2–6) |
|  | >1 symptom | 6 (4–9) | 4 (3–6) |
| AGE symptoms |  |  |  |
|  | Yes | 7 (3–12) | 3 (2–5) |
|  | No | 5 (3–9) | 4 (2–6) |

**Supplementary figure 1.**
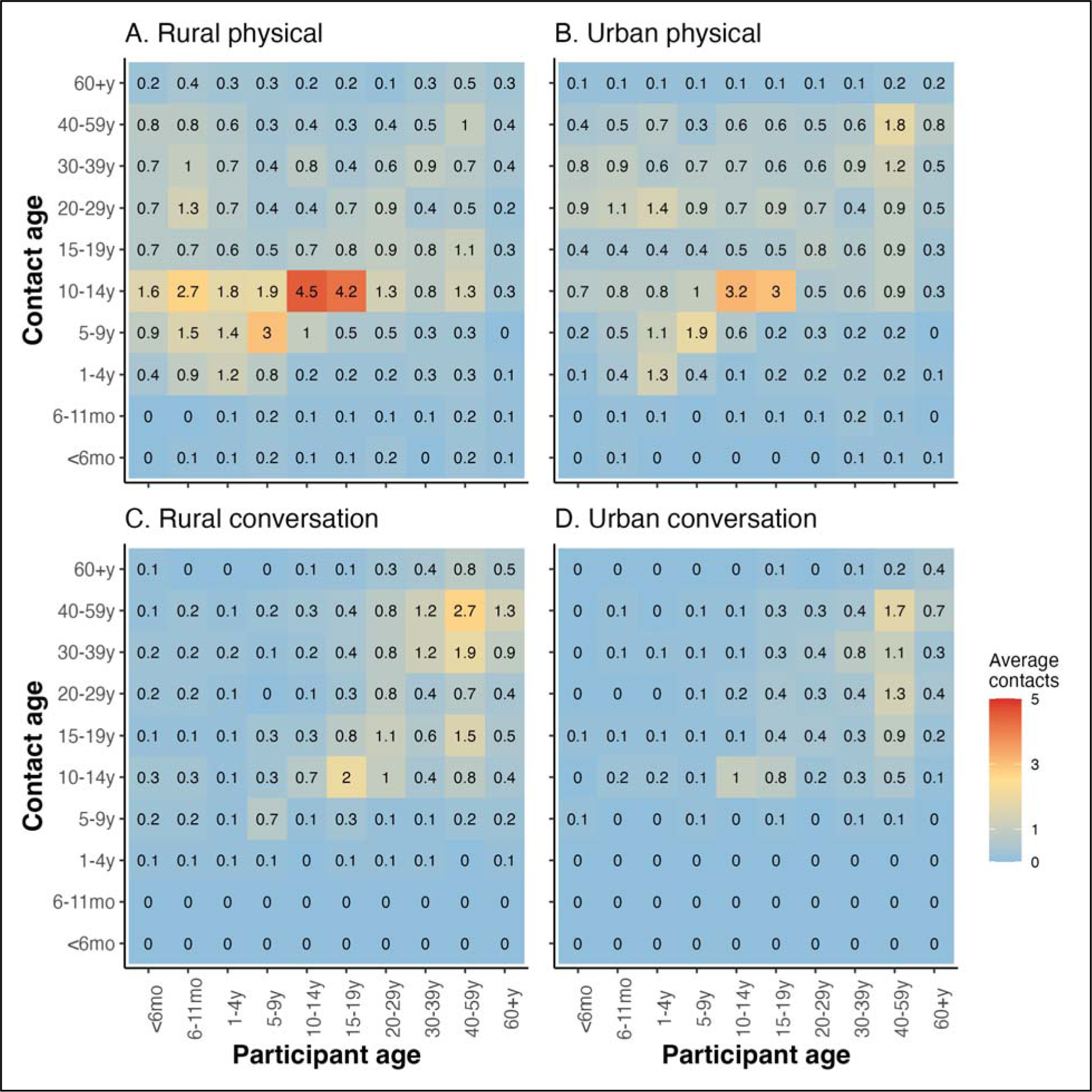
Matrices of age–specific average physical only and conversation only contacts by site. The panels are arranged by rural (A) and urban (B) physical contacts and rural (C) and urban (D) conversation contacts. Average age-specific values are generated by dividing the total number of contacts reported by age group pair with the number of participants in that age group.

**Supplementary figure 2.**
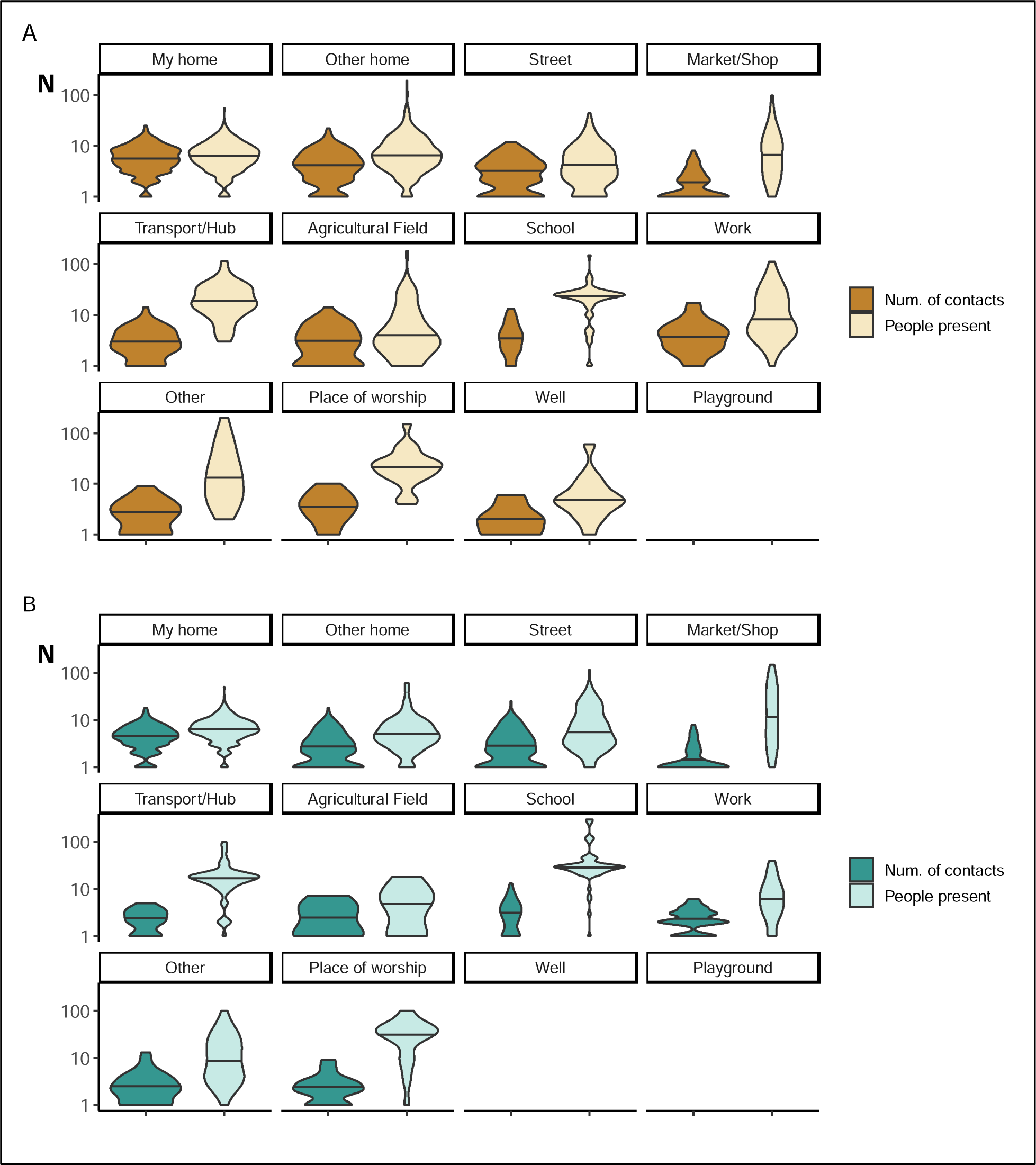
Violin plots comparing the distribution of number of contacts versus number of co-located individuals reported by participants at each location in rural (A) versus urban (B) site.

